# Trajectories of Estimated Glucose Disposal Rate and Risk of Cardiovascular Disease: Evidence from Two Prospective Cohorts

**DOI:** 10.64898/2026.11.02.26367136

**Authors:** Rong-Shuai Pan, Xicong Li, Zi-Hang Wei, Jie Zou, Ze-Ru Chen, Xi Liang, Wen-Kai He, Li Mai, Ming-Yan Li

## Abstract

**Background:** Estimated glucose disposal rate (eGDR) is an accessible surrogate of insulin sensitivity and has been associated with cardiovascular disease (CVD) risk. However, whether longitudinal eGDR trajectory patterns convey prognostic information beyond cumulative eGDR exposure remains unclear. We aimed to characterize distinct eGDR trajectories and evaluate their associations with incident CVD in two prospective cohorts.

**Methods:** We included 4,823 CVD-free adults aged 45 years or older from the Health and Retirement Study (HRS, n=3,258) and the English Longitudinal Study of Ageing (ELSA, n=1,565). Latent class mixed models identified four eGDR trajectory classes across three waves. Associations with incident CVD were examined using multivariable Cox models. We further fitted joint models including both trajectory class and cumulative average eGDR to assess their independent contributions.

**Results:** Compared with the persistently high trajectory, the persistently low and declining trajectories were associated with higher CVD risk in both cohorts. In joint models, whereas trajectory class showed residual prognostic signal, particularly in HRS, with directionally consistent but less precise estimates in ELSA. In HRS, adding trajectory class improved model fit beyond cumulative average eGDR alone, whereas adding cumulative average eGDR to the trajectory model did not. Direct pairwise comparisons showed that the declining-then-rising trajectory was associated with lower risk than the declining trajectory in HRS, with directionally similar but imprecise estimates in ELSA.

**Conclusions:** Longitudinal eGDR trajectory patterns may convey prognostic information not fully captured by cumulative average eGDR alone. These findings suggest that the temporal pattern of insulin sensitivity may be clinically relevant in addition to cumulative exposure, while the potentially favorable profile of recovery trajectories warrants cautious interpretation and external replication.

**Graphical abstract:** 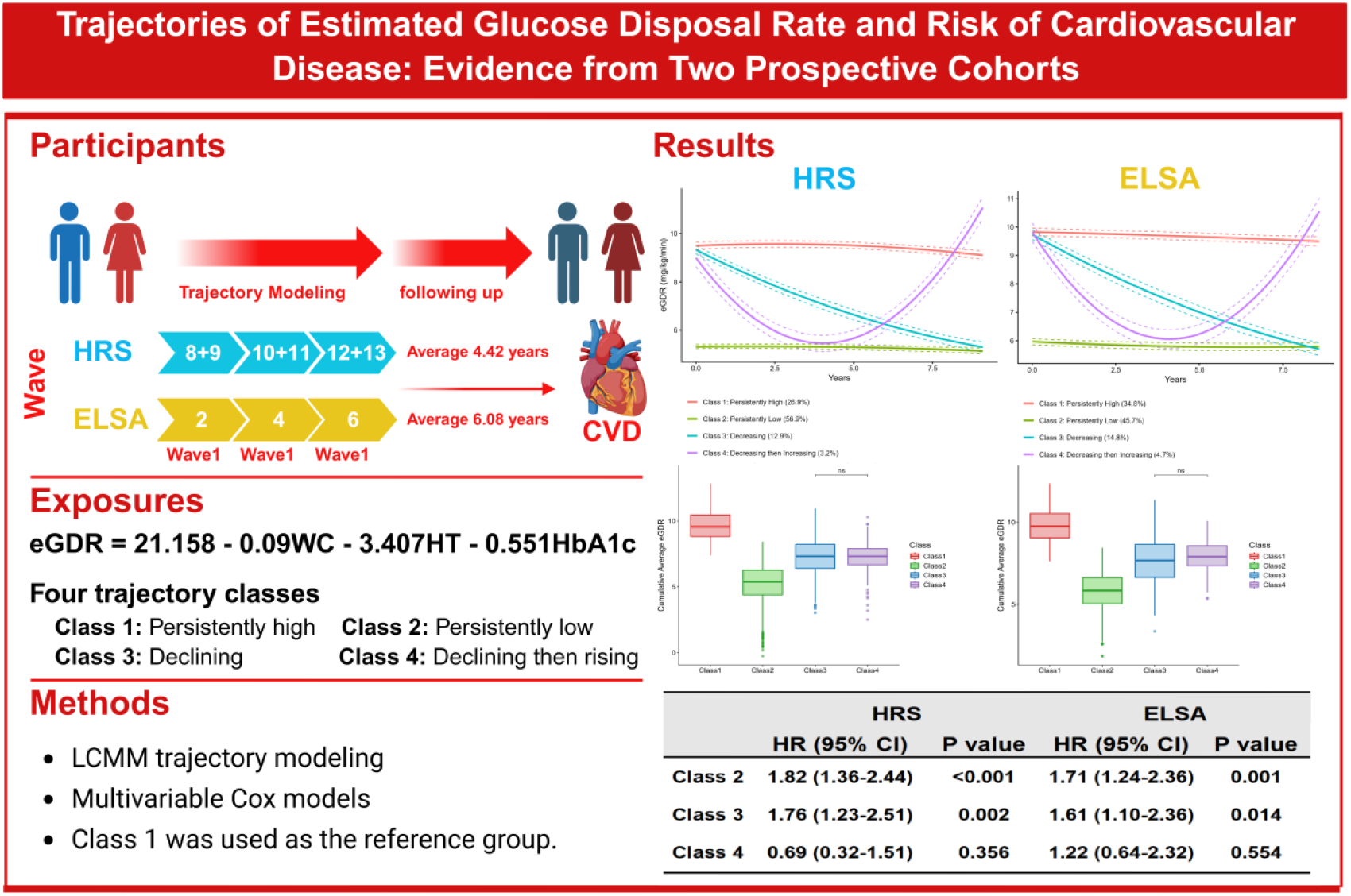

## Introduction

Cardiovascular disease (CVD) remains the leading cause of morbidity and premature mortality worldwide, imposing a substantial burden on healthcare systems and public health.(1–3) Despite substantial advances in diagnosis and treatment over recent decades, the global prevalence of CVD continues to rise, largely driven by population aging and the increasing prevalence of metabolic diseases.(4,5) Early identification of modifiable risk factors and individuals at high risk has therefore become a priority for reducing the burden of CVD.

Insulin resistance (IR), a pathological state characterized by reduced cellular responsiveness to insulin, is central to metabolic dysregulation and constitutes a key mechanistic link between metabolic syndrome, type 2 diabetes mellitus, and atherosclerotic cardiovascular disease(6,7). IR promotes a pro-inflammatory state, endothelial dysfunction, and dyslipidemia, thereby accelerating vascular injury even in the absence of overt hyperglycemia(8–11). Although the hyperinsulinemic-euglycemic clamp technique is considered the gold standard for quantifying insulin resistance, its technical complexity, high cost, and time-consuming nature preclude its large-scale application in epidemiological studies or routine clinical screening.(12,13) Similarly, the homeostatic model assessment of insulin resistance (HOMA-IR) is widely used but requires fasting insulin measurements, which are not routinely collected in many population-based cohorts(14–17), thereby limiting its applicability in large-scale research.

To address these limitations, surrogate markers including the triglyceride-glucose (TyG) index and the estimated glucose disposal rate (eGDR) have been developed. The eGDR, calculated from waist circumference, hypertension status, and glycated hemoglobin (HbA1c)(18), has emerged as a reliable and clinically feasible index of insulin resistance(19). A growing body of evidence suggests that eGDR may outperform other surrogate indices, including TyG, in predicting adverse outcomes such as diabetes, all-cause mortality, and CVD(20–24). Lower eGDR values, reflecting greater insulin resistance, have been independently associated with increased risks of stroke, coronary heart disease, and cardiovascular death across diverse populations(20,24).

However, most existing studies have relied on single baseline measurements of eGDR to predict CVD risk.(22,25–27) This static approach fails to capture the dynamic nature of metabolic health and overlooks the potential implications of longitudinal changes in insulin sensitivity. Latent class mixed modeling (LCMM) is a widely used trajectory analysis approach that can simultaneously capture individual-level longitudinal change and population-level heterogeneity, and has been extensively applied in medical and epidemiological research.(28,29) Recent studies using trajectory modeling of other insulin resistance surrogates, such as HOMA-IR and TyG, have shown that distinct longitudinal patterns confer differential risks of CVD incidence and mortality, often with greater prognostic value than baseline measurements alone.(30–37) In particular, deteriorating insulin resistance trajectories have been associated with substantially elevated cardiovascular risk, underscoring the importance of monitoring temporal trends. At the same time, cumulative exposure to cardiometabolic risk has also been linked to future cardiovascular outcomes, and recent studies have begun to evaluate cumulative burden and dynamic trajectories as potentially complementary longitudinal dimensions of risk.(37,38)

Despite these advances, no study has specifically examined longitudinal eGDR trajectories in relation to incident CVD. In the present study, we used latent class mixed models to identify distinct eGDR trajectories in two large prospective cohorts of middle-aged and older adults—the Health and Retirement Study (HRS) in the United States and the English Longitudinal Study of Ageing (ELSA) in England—and examined their associations with incident CVD. We further tested whether trajectory class provides prognostic information beyond cumulative average eGDR.

Taken together, these findings suggest that cumulative exposure and trajectory pattern may be related but not fully interchangeable. Cumulative average eGDR reflects the overall long-term metabolic burden, whereas trajectory class captures the temporal pattern of change— two related but potentially non-identical dimensions of longitudinal exposure. Individuals with similar cumulative average eGDR may therefore still follow different metabolic courses and potentially experience different cardiovascular consequences. We therefore hypothesized that eGDR trajectory class and cumulative average eGDR might provide overlapping but non- identical prognostic information for incident CVD.

## Methods

### Study Design and Participants

This study utilized data from two internationally harmonized, population-based longitudinal cohort studies: the Health and Retirement Study (HRS) conducted in the United States and the English Longitudinal Study of Ageing (ELSA) conducted in England. The HRS is a nationally representative biennial panel survey of community-dwelling Americans aged ≥50 years, established in 1992 and maintained by the University of Michigan; biomarker data from dried blood spots were available across six waves spanning 2006–2016(39). ELSA is a representative longitudinal cohort of community-dwelling adults aged ≥50 years in England, established in 2002; fasting venous blood samples were obtained during dedicated nurse visit waves (waves 2, 4, and 6, corresponding to approximately 2004–2005, 2008–2009, and 2012–2013, respectively). (40)

Participants were eligible for inclusion if they: (1) were aged ≥45 years at baseline; (2) had no prevalent CVD prior to the end of the exposure ascertainment period; (3) had complete data for all three eGDR component variables (waist circumference, hypertension status, and HbA1c) across three consecutive waves; and (4) had eGDR values within three standard deviations (SDs) of the cohort-specific mean at each wave. Participants with extreme outlier values beyond this threshold were excluded to minimize measurement error. After applying these criteria, the analytic cohort comprised 4,823 participants (HRS: n = 3,258; ELSA: n = 1,565). The study inclusion and exclusion process is summarized in Supplementary Figure S1.

The HRS was approved by the University of Michigan Health Sciences and Behavioral Sciences Institutional Review Board (Protocol No. HUM00061128). ELSA received ethical approval from the relevant UK Research Ethics Committees across waves (Wave 2: MREC/04/2/006; Wave 4: 07/H0716/48; Wave 6: 11/SC/0374). All participants provided written informed consent. The present study involved secondary analysis of de-identified publicly available data and required no additional institutional ethical review.

### Assessment of the change in eGDR

The estimated glucose disposal rate (eGDR), was calculated using the following formula(24) eGDR (mg/kg/min)=21.158-(0.09×WC)-(3.407×HT)-(0.551×HbA1c) where WC denotes waist circumference (cm), HT represents hypertension status (1 = yes, 0 = no), and HbA1c indicates glycated haemoglobin (%). Hypertension was defined through self-reported physician diagnosis of hypertension, measured systolic blood pressure ≥140 mmHg or diastolic blood pressure ≥90 mmHg, or current use of antihypertensive medication.(41) In ELSA wave 6, HbA1c was measured using the International Federation of Clinical Chemistry (IFCC) standard and converted to the National Glycohemoglobin Standardization Program (NGSP) equivalent using the formula(42): HbA1c^NGSP^ = 0.0915 × HbA1c^IFCC^ + 2.15,prior to eGDR calculation, ensuring comparability across cohorts.

Five longitudinal metrics were derived from the three serial eGDR values(43): (1) cumulative eGDR (trapezoidal integration: Σ [(eGDRi + eGDRi+1) / 2 × (ti+1 − ti)]); (2) cumulative average eGDR (cumulative eGDR divided by total follow-up years); (3) annual slope ((eGDRlast − eGDRfirst) / years); (4) visit-to-visit variability (SD, CV, and VIM = 100 × SD / meanβ, where β was estimated by log-log regression); and (5) trajectory class (described below).

### Trajectory Modeling

Latent class mixed models (LCMM) were fitted separately in each cohort using the lcmm R package(44) to identify subgroups with distinct eGDR trajectories. Grid search with 30 random starting values was applied to avoid local optima. Models with 2–4 classes were evaluated; the optimal solution was selected based on minimum BIC/AIC/aBIC, entropy ≥0.70, and minimum class membership ≥3%.(45–48) A four-class solution was selected in both cohorts. Each participant was assigned to the class with the highest posterior probability.

### Outcome

The primary outcome was incident cardiovascular disease (CVD), defined as new-onset heart disease or stroke. In both the HRS and ELSA, CVD events were ascertained through self-reported physician-diagnosed conditions using a standardized question battery administered at each wave. Specifically, participants were asked: “Has a doctor ever told you that you have been diagnosed with a heart disease (including myocardial infarction, coronary heart disease, angina, congestive heart failure, or other heart problems)?” and “Has a doctor ever told you that you have been diagnosed with a stroke?”. Participants who answered affirmatively to either question at a follow-up wave were classified as having incident CVD. This definition is consistent with previous large-scale studies using these cohorts.(49,50) Each cohort employs slightly different variable names for these self-reported items. A detailed mapping of the survey variables used to ascertain heart disease and stroke across waves is provided in Supplementary Table S12. For interval-censored events, the midpoint between the wave of first positive report and the preceding wave was used as the event date. Participants who remained free of CVD were censored at the date of the last completed interview, death, or end of follow-up, whichever occurred first. Follow-up began after the final eGDR assessment wave (wave 3 exposure period) and extended through the most recent available outcome wave.(25)

### Covariates and Missing Data

Baseline covariates included age, sex, education, marital status, smoking, alcohol use, BMI, Physical activity, history of diabetes/cancer/lung disease, antidiabetic and lipid-lowering medication use, and biomarkers (HbA1c, total cholesterol, HDL-C, CRP). Waist circumference, blood pressure were excluded from adjustment to avoid over-adjustment. Analyses of eGDR change metrics additionally adjusted for baseline eGDR(25,51).

Missing covariates were handled by multiple imputation by chained equations (MICE; m = 5, 50 iterations), applying predictive mean matching for continuous variables, logistic regression for binary variables, and polytomous regression for nominal variables, performed separately per cohort.

### Statistical Analysis

Baseline characteristics were compared across the four eGDR trajectory classes. Continuous variables were assessed for normality and presented as mean ± standard deviation or median (interquartile range), as appropriate. Differences across trajectory classes were tested using one-way analysis of variance for normally distributed variables or the Kruskal–Wallis test for skewed variables. Categorical variables were presented as counts (percentages) and compared using chi-square tests or Fisher’s exact tests, as appropriate. These descriptive results are presented in Supplementary Tables S1 and S2 for HRS and ELSA, respectively. Details of trajectory model selection are provided in Supplementary Table S3.

Associations between eGDR metrics and incident CVD were evaluated using multivariable Cox proportional hazards regression across three prespecified nested models. Model 1 adjusted for age and sex. Model 2 additionally adjusted for education, marital status, smoking status, alcohol consumption, body mass index, and physical activity. Model 3 further adjusted for comorbidities (history of diabetes, cancer, and lung disease), medication use (antidiabetic and lipid-lowering drugs), and biomarkers (HbA1c, total cholesterol, HDL-C, and C-reactive protein). Trajectory class and cumulative average eGDR were first examined separately. We then fitted joint Cox models including both trajectory class and cumulative average eGDR to assess their independent contributions. To further evaluate whether the declining-then-rising trajectory carried a distinct risk profile, direct pairwise comparisons were performed for Class 4 versus Class 3 and for Class 4 versus Class 2.

Restricted cubic spline models with three knots were fitted using the rms package to examine the dose-response relationship between cumulative average eGDR and CVD. Prediction analyses, including Harrell’s C-index and time-dependent area under the receiver operating characteristic curve at 1, 3, and 5 years, were treated as supportive rather than primary analyses. Subgroup analyses were performed by age, sex, smoking status, and BMI, with interactions tested using likelihood ratio tests. Sensitivity analyses included exclusion of participants with prevalent diabetes at baseline, complete-case analysis to reduce potential reverse causality, and a three-class trajectory solution. All tests were two-sided, and P<0.05 was considered statistically significant. All analyses were performed using R version 4.5.0, with survival, rms, timeROC, and lcmm as the core packages.

## Results

### Baseline Characteristics

A total of 4,823 participants were included in this analysis, drawn from the HRS cohort (N = 3,258) and the ELSA cohort (N = 1,565). Overall, the median age of HRS participants was 64.00 (Q1–Q3: 57.00–70.00) years, and the median age of ELSA participants was 61.00 (56.00–67.00) years, as detailed in Supplementary Table S1 and S2.

### Identification of eGDR Trajectories

Based on LCMM, four distinct eGDR trajectories were identified in both cohorts (Figure 1A and 1B). The optimal four-class solution was determined based on the lowest BIC/AIC/aBIC values, mean posterior probabilities >0.70 for each class, a minimum class proportion ≥3%, and clinical interpretability(Table S3). The four trajectory classes were as follows: Class 1, persistently high (HRS: 26.9%; ELSA: 34.8%); Class 2, persistently low (HRS: 56.9%; ELSA: 45.7%); Class 3, declining (HRS: 12.9%; ELSA: 14.8%); and Class 4, declining then rising (HRS: 3.2%; ELSA: 4.7%). Although Class 4 accounted for a relatively small proportion of participants, it was retained because it represented a clinically distinct recovery pattern and included more than 50 participants in each cohort. The corresponding three-class solution was examined as a sensitivity analysis rather than as the primary model. Figure 1C and 1D illustrate the distribution of cumulative average eGDR across trajectory classes.

**Figure 1.**
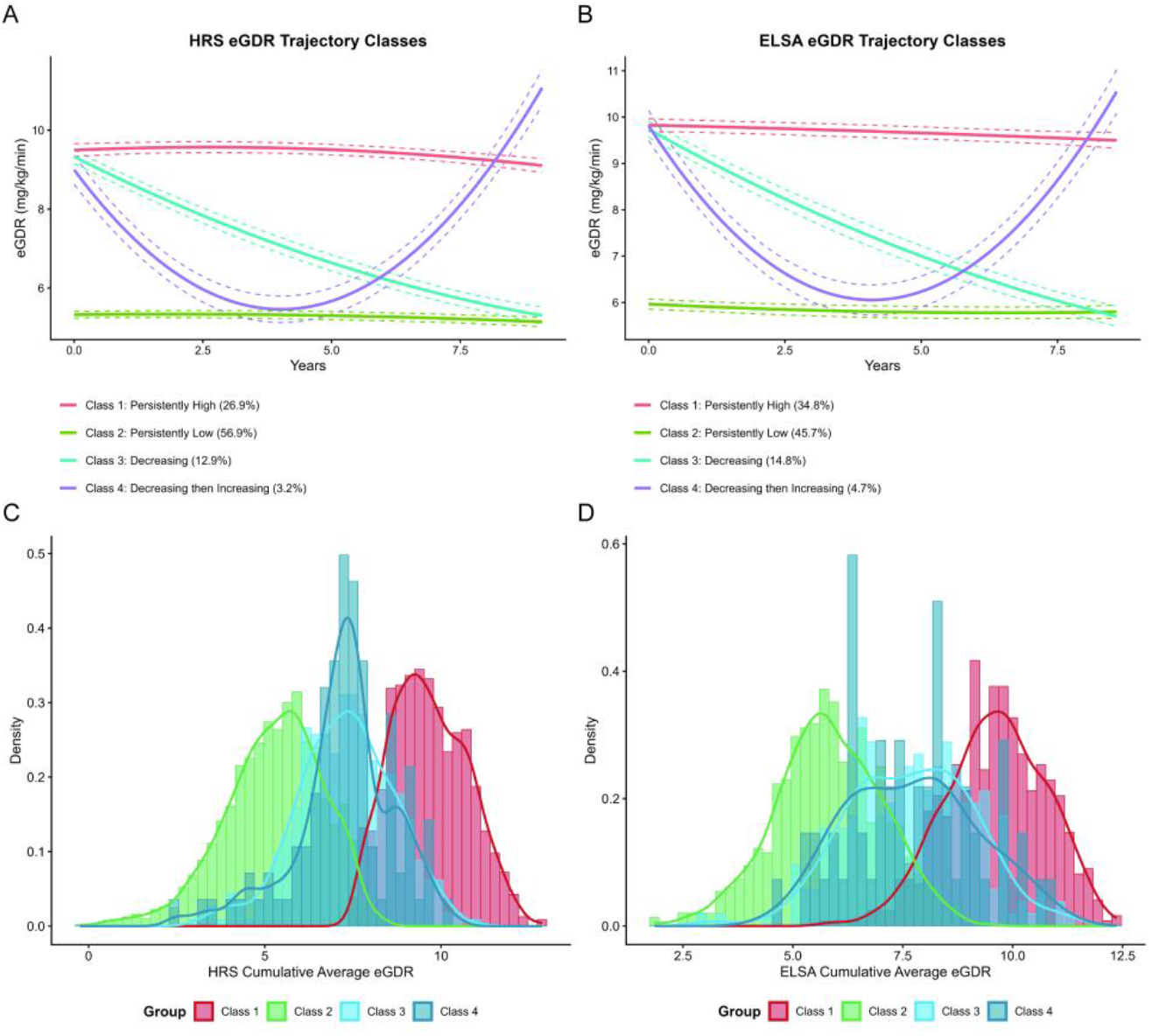
eGDR trajectory classes and distribution of cumulative average eGDR in the HRS and ELSA cohorts.

### Association Between eGDR Trajectories and CVD Risk

During the trajectory assessment phase, the observation window spanned a median of 7.83 years (IQR, 7.67–8.00) in ELSA and 8.01 years (IQR, 7.84–8.17) in HRS. During the subsequent outcome surveillance phase, 253 incident CVD events (16.17%) were recorded in ELSA over a median follow-up of 6.08 years (IQR, 4.17–6.17), and 416 incident CVD events (12.77%) were recorded in HRS over a median follow-up of 4.42 years (IQR, 3.83–5.92).

Table 1 presents the associations of trajectory class and cumulative average eGDR with incident CVD. In separate multivariable Cox models, both trajectory class and cumulative average eGDR were associated with incident CVD. Compared with Class 1 (persistently high), Class 2 (persistently low) was associated with higher CVD risk in both HRS (HR 1.82, 95% CI 1.36–2.44; P<0.001) and ELSA (HR 1.71, 95% CI 1.24–2.36; P=0.001). Class 3 (declining) was also associated with elevated risk in HRS (HR 1.76, 95% CI 1.23–2.51; P=0.002) and ELSA (HR 1.61, 95% CI 1.10–2.36; P=0.014). In contrast, Class 4 (declining-then-rising) was not significantly different from Class 1 in either cohort.

**Table 1.** Association of eGDR trajectory class and cumulative average eGDR with incident cardiovascular disease.

| **Variable** | **CVD Case, n (%)** | **Model 1** |  | **Model 2** |  | **Model 3** |  |
|---|---|---|---|---|---|---|---|
|  |  | **HR (95% CI)** | **P value** | **HR (95% CI)** | **P value** | **HR (95% CI)** | **P value** |
| **HRS, N = 3258** |  |  |  |  |  |  |  |
| **eGDR trajectory class** |  |  |  |  |  |  |  |
| Class 1 | 62 (7.06%) | Ref |  | Ref |  | Ref |  |
| Class 2 | 295 (15.91%) | 2.04 (1.56-2.67) | < 0.001 | 1.91 (1.43-2.54) | < 0.001 | 1.82 (1.36-2.44) | < 0.001 |
| Class 3 | 53 (12.59%) | 1.81 (1.27-2.58) | < 0.001 | 1.77 (1.24-2.52) | 0.002 | 1.76 (1.23-2.51) | 0.002 |
| Class 4 | 6 (5.71%) | 0.74 (0.34-1.61) | 0.449 | 0.71 (0.33-1.56) | 0.399 | 0.69 (0.32-1.51) | 0.356 |
| **Cumulative average eGDR** |  | 0.88 (0.84-0.92) | < 0.001 | 0.87 (0.82-0.93) | < 0.001 | 0.88 (0.83-0.94) | < 0.001 |
| **Quartiles** |  |  |  |  |  |  |  |
| Q1(<5.10) | 136 (16.69%) | Ref |  | Ref |  | Ref |  |
| Q2(5.10-6.49) | 114 (14.00%) | 0.80 (0.63-1.02) | 0.077 | 0.83 (0.64-1.08) | 0.174 | 0.89 (0.68-1.18) | 0.431 |
| Q3(6.49-8.59) | 113 (13.88%) | 0.80 (0.63-1.02) | 0.073 | 0.84 (0.63-1.12) | 0.233 | 0.92 (0.68-1.24) | 0.567 |
| Q4(≥8.59) | 53 (6.50%) | 0.42 (0.31-0.57) | < 0.001 | 0.44 (0.31-0.64) | < 0.001 | 0.50 (0.34-0.74) | < 0.001 |
| **P for trend** |  |  | < 0.001 |  | < 0.001 |  | 0.003 |
| **ELSA, N = 1,565** |  |  |  |  |  |  |  |
| **eGDR trajectory class** |  |  |  |  |  |  |  |
| Class 1 | 60 (11.01%) | Ref |  | Ref |  | Ref |  |
| Class 2 | 137 (19.16%) | 1.66 (1.23-2.24) | < 0.001 | 1.73 (1.25-2.38) | < 0.001 | 1.71 (1.24-2.36) | 0.001 |
| Class 3 | 45 (19.40%) | 1.66 (1.14-2.43) | 0.008 | 1.67 (1.14-2.44) | 0.009 | 1.61 (1.10-2.36) | 0.014 |
| Class 4 | 11 (15.07%) | 1.22 (0.64-2.31) | 0.546 | 1.19 (0.62-2.26) | 0.598 | 1.22 (0.64-2.32) | 0.554 |
| **Cumulative average eGDR** |  | 0.91 (0.86-0.97) | 0.003 | 0.89 (0.82-0.96) | 0.002 | 0.89 (0.82-0.96) | 0.004 |
| **Quartiles** |  |  |  |  |  |  |  |
| Q1(<5.86) | 68 (17.35%) | Ref |  | Ref |  | Ref |  |
| Q2(5.86-7.48) | 83 (21.23%) | 1.23 (0.90-1.68) | 0.194 | 1.20 (0.85-1.69) | 0.297 | 1.18 (0.83-1.69) | 0.353 |
| Q3(7.48-9.27) | 66 (16.88%) | 0.93 (0.67-1.30) | 0.682 | 0.88 (0.61-1.27) | 0.488 | 0.89 (0.62-1.30) | 0.559 |
| Q4(≥9.27) | 36 (9.21%) | 0.57 (0.38-0.85) | 0.005 | 0.54 (0.34-0.86) | 0.009 | 0.55 (0.34-0.88) | 0.013 |
| **P for trend** |  |  | 0.004 |  | 0.003 |  | 0.004 |

Note: Independent Contributions of Trajectory Class and Cumulative Average eGDR. Model 1 adjusted for age, sex; Model 2 further adjusted for education level, marital status, smoking status, drinking status BMI and physical activity based on model 1; Model 3 further adjusted for history of disease,medicine and biomarkers based on model 2

Cumulative average eGDR was inversely associated with incident CVD in both cohorts. Each 1-SD increment in cumulative average eGDR was associated with a 12% lower CVD risk in HRS (HR 0.88, 95% CI 0.83–0.94; P<0.001) and an 11% lower risk in ELSA (HR 0.89, 95% CI 0.82–0.96; P=0.004). Quartile analyses showed a graded reduction in CVD risk with increasing cumulative average eGDR (P for trend=0.003 in HRS and 0.004 in ELSA), with the highest quartile associated with a 50% lower risk than the lowest quartile in HRS (HR 0.50, 95% CI 0.34–0.74) and a 45% lower risk in ELSA (HR 0.55, 95% CI 0.34–0.88).

Notably, despite similar distributions of cumulative average eGDR between Classes 3 and 4 (Supplementary Figure S2), their CVD risk outcomes differed, suggesting that eGDR trajectory shape may reflect prognostic information not fully interchangeable with total exposure burden. Among these metrics, trajectory class and cumulative average eGDR were prespecified as the two primary longitudinal summaries of interest, whereas slope and variability indices were treated as secondary descriptors of temporal change (Table S4).

To further characterize the association between cumulative average eGDR and incident CVD, restricted cubic spline analyses were performed (Figure 2). After adjustment for Model 3 covariates, cumulative average eGDR showed an approximately linear inverse association with CVD risk in both cohorts, with significant overall associations in HRS (overall P<0.001; nonlinearity P=0.128) and ELSA (overall P=0.004; nonlinearity P=0.092). These findings were consistent with the quartile analyses and support an inverse association between sustained higher cumulative average eGDR and lower CVD risk. Corresponding spline analyses for heart disease and stroke as secondary outcomes are presented in Supplementary Figure S3.

**Figure 2.**
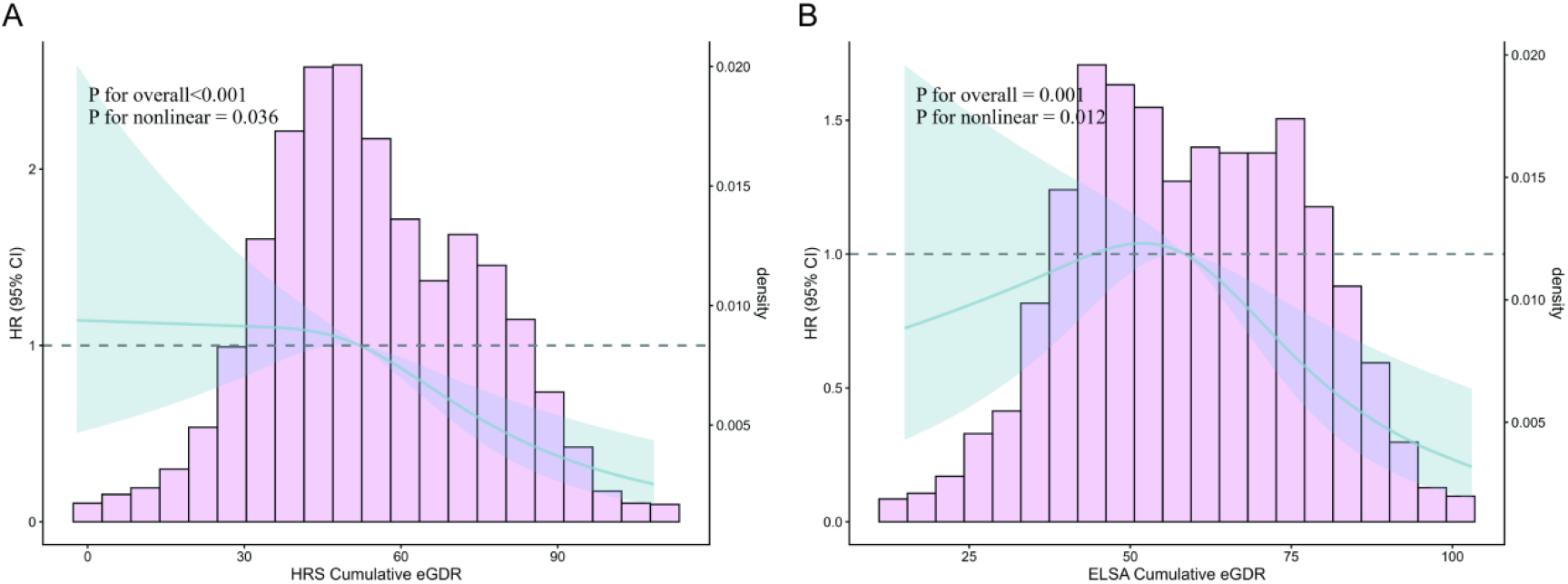
Restricted cubic spline curves for cardiovascular disease according to cumulative average eGDR in the HRS and ELSA cohorts. Cox models were adjusted for the same covariates as in Model 3 of Table 1.

Analyses of annual eGDR change are shown in Supplementary Figure S4. A linear association between annual eGDR change and incident CVD was observed in HRS (overall P=0.002; nonlinearity P=0.507), whereas no statistically significant association was detected in ELSA (overall P=0.271; nonlinearity P=0.871).

### Independent Contributions of Trajectory Class and Cumulative Average eGDR

To evaluate whether trajectory class and cumulative average eGDR provided independent prognostic information, we fitted joint Cox models including both variables simultaneously (Table 2). In HRS, cumulative average eGDR was no longer associated with incident CVD after adjustment for trajectory class (HR 0.959, 95% CI 0.869–1.058; P=0.404), whereas Class 2 and Class 3 remained associated with higher risk relative to Class 1 (Class 2: HR 1.589, 95% CI 1.029–2.455; P=0.037; Class 3: HR 1.620, 95% CI 1.079– 2.432; P=0.020). In ELSA, cumulative average eGDR was likewise not independently associated with incident CVD (HR 0.978, 95% CI 0.849–1.127; P=0.760), and the estimates for Class 2 and Class 3 remained directionally consistent with those in HRS, although with wider confidence intervals and without statistical significance. Class 4 was not significantly different from Class 1 in either cohort.

**Table 2.** Joint Cox models and direct pairwise comparisons of eGDR trajectory classes and cumulative average eGDR with incident cardiovascular disease.

|  | **Variable** | **HRS** |  | **ELSA** |  |
|---|---|---|---|---|---|
|  |  | **HR (95% CI)** | **P value** | **HR (95% CI)** | **P value** |
| **Panel A**<br>**Joint Cox model** | Class 2 vs Class 1<br>(persistently low vs persistently high) | 1.589 (1.029-2.455) | 0.037 | 1.595 (0.910-2.794) | 0.103 |
|  | Class 3 vs Class 1<br>(declining vs persistently high) | 1.620 (1.079-2.432) | 0.020 | 1.549 (0.976-2.460) | 0.063 |
|  | Class 4 vs Class 1<br>(declining-then-rising vs persistently high) | 0.640 (0.286-1.431) | 0.277 | 1.169 (0.585-2.335) | 0.659 |
|  | Cumulative average eGDR<br>(per 1-unit increase) | 0.959 (0.869-1.058) | 0.404 | 0.978 (0.849-1.127) | 0.760 |
| **Panel B.**<br>**Direct pairwise comparisons** | Class 4 vs Class 2<br>(recovering vs persistently low) | 0.383 (0.180-0.814) | 0.013 | 0.680 (0.360-1.283) | 0.234 |
|  | Class 4 vs Class 3<br>(recovering vs declining) | 0.398 (0.179-0.884) | 0.024 | 0.780 (0.393-1.547) | 0.476 |
| **Panel C**<br>**Incremental model fit** | Incremental fit: add cumulative average eGDR beyond trajectory class | LRT P = 0.402 |  | LRT P = 0.759 |  |
|  | Incremental fit: add trajectory class beyond cumulative average eGDR | LRT P = 0.011 |  | LRT P = 0.286 |  |

**Note:** Panel A presents mutual-adjustment Cox models including both trajectory class and cumulative average eGDR. Panel B presents direct comparisons for the recovering trajectory (Class 4) against metabolically adverse trajectories. Panel C presents likelihood ratio tests for incremental model fit. Model adjustment: age, sex, education, marital status, smoking, drinking, BMI, physical activity, history of diabetes/cancer/lung disease, antidiabetic and lipid-lowering medication use, and cohort-specific biomarkers.

Direct pairwise comparisons were then performed to evaluate the recovering trajectory against metabolically adverse trajectories. In HRS, Class 4 was associated with lower CVD risk than both Class 2 (HR 0.383, 95% CI 0.180–0.814; P=0.013) and Class 3 (HR 0.398, 95% CI 0.179–0.884; P=0.024). In ELSA, the corresponding estimates were directionally similar but not statistically significant.These ELSA estimates should be interpreted cautiously given the wider confidence intervals and smaller cohort size. Likelihood ratio tests further showed that, in HRS, adding trajectory class significantly improved model fit beyond cumulative average eGDR alone (P=0.011), whereas adding cumulative average eGDR to the trajectory model did not (P=0.402). In ELSA, neither incremental comparison reached statistical significance, but cumulative average eGDR again showed no evidence of material added value once trajectory class was known,This is very likely caused by an insufficient sample size.

Supportive prediction analyses showed that models incorporating trajectory class generally performed as well as or better than models based on cumulative average eGDR alone across the evaluated prediction horizons. However, the absolute improvements in discrimination were modest, and these analyses were considered secondary to the etiologic interpretation of the joint Cox models. Detailed results for time-dependent AUC, Harrell’s C- index, calibration, decision-curve analyses, and secondary-outcome prediction for heart disease and stroke are presented in the Supplementary Material(Supplementary Figure S5- S11, Table S5-S8).

### Subgroup Analyses

Subgroup analyses stratified by age, sex, smoking status, and BMI are presented in Supplementary Figures S12 and S13. The elevated CVD risk associated with adverse trajectory classes and the lower risk associated with higher cumulative average eGDR (Q4 vs Q1) were broadly consistent across subgroups in both cohorts. No statistically significant interactions were detected for trajectory class or cumulative average eGDR (all P for interaction >0.05), suggesting that the primary associations were generally stable across the examined participant characteristics.

### Sensitivity Analyses

Several sensitivity analyses were conducted to assess the robustness of the primary findings. (1) Exclusion of participants with baseline diabetes and complete-case analyses without multiple imputation yielded results broadly consistent with the primary analysis (Tables S8 and S9). (2)The three-class trajectory solution was used as a robustness check and showed broadly similar associations with incident CVD in both cohorts (Supplementary Figure S14 and Table S11). However, because the declining and recovering patterns were not separated in the three-class solution, this analysis could not evaluate whether the declining- then-rising trajectory carried a distinct risk profile.

### Discussion

In this study, we used LCMM to characterize longitudinal eGDR trajectories and examined their associations with incident CVD in two large, internationally harmonized prospective cohorts of middle-aged and older adults. We found that eGDR trajectory class and cumulative average eGDR were not interchangeable. Although both measures were associated with incident CVD when examined separately, cumulative average eGDR was no longer independently associated with CVD once trajectory class was taken into account, whereas the persistently low and declining trajectory classes retained prognostic signal, particularly in HRS. These findings suggest that the temporal pattern of insulin sensitivity may convey clinically relevant information beyond average long-term exposure alone.

Several previous studies have confirmed that lower baseline eGDR was independently associated with CVD events. Dong et al. reported that each unit increment in eGDR was associated with a 14% reduction in CVD risk among individuals with cardiovascular-kidney- metabolic syndrome stages 0–3, with eGDR outperforming six other insulin resistance indices.(52) Zhang et al. similarly found that each SD increment in eGDR corresponded to a 17% lower CVD risk in a nationwide non-diabetic cohort(14). A recent systematic review and meta-analysis further confirmed the robust predictive value of eGDR across diverse populations.(53) Our findings are consistent with this body of evidence and further suggest that longitudinal trajectory pattern may convey prognostic information not fully captured by time- averaged exposure, though the strength and consistency of this observation varied between cohorts.

While these studies established the prognostic value of single-timepoint eGDR, research on its dynamic trajectories remains limited. Recent study examined eGDR changes between two timepoints using K-means clustering and found that both persistently low and low-rising eGDR patterns were associated with elevated CVD risk. (25,51) However, approaches based on only two measurements cannot fully capture intra-individual variation over time or the continuity of trajectory evolution. Likewise, cumulative eGDR exposure reflects overall metabolic burden, but does not distinguish whether similar long-term exposure arose from persistently impaired insulin sensitivity, progressive deterioration, or partial recovery. By applying LCMM and evaluating trajectory class alongside cumulative average eGDR within the same analytic framework, our study helps clarify that these two longitudinal summaries are related but not equivalent. This approach has similarly shown that deteriorating longitudinal patterns carry excess cardiovascular risk beyond baseline values alone(30–37). We also introduced cumulative average eGDR as a refined metric that corrects for variable follow-up intervals, providing a more reliable measure of sustained insulin sensitivity exposure.

A particularly noteworthy finding was the divergence in CVD risk between Class 3 (declining) and Class 4 (declining-then-rising) trajectories, despite their broadly comparable cumulative average eGDR distributions. This distinction is clinically relevant because individuals with similar cumulative average eGDR may still follow biologically different metabolic courses and experience different vascular consequences. In this sense, trajectory class is not merely a more complicated representation of cumulative exposure, but may reflect a distinct dimension of cardiometabolic risk.

The declining-then-rising trajectory deserves cautious interpretation. In HRS, direct pairwise comparisons showed that this trajectory was associated with lower CVD risk than both the persistently low and declining trajectories, raising the hypothesis that recovery after decline could carry prognostic meaning, pending replication in larger studies. However, the corresponding estimates in ELSA were directionally similar but imprecise and did not reach statistical significance, and Class 4 was not significantly different from Class 1 in either cohort. Accordingly, the potentially favorable profile of the declining-then-rising trajectory should be regarded as hypothesis-generating rather than definitive evidence of cardiovascular protection. One possible explanation is that recovery after decline may reflect partial restoration of endothelial function, attenuation of inflammatory burden, or favorable behavioral or therapeutic modification, but these mechanisms cannot be confirmed in the current study.(10,11,54)

From a prediction perspective, models incorporating trajectory class generally performed as well as or better than models based on cumulative average eGDR alone, although the absolute improvements in discrimination were modest. We therefore consider the predictive analyses supportive rather than central to the main etiologic interpretation. Given that eGDR can be calculated from routinely available clinical parameters, repeated assessment may still offer a practical way to monitor longitudinal cardiometabolic risk in middle-aged and older adults.

Subgroup analyses showed broadly consistent associations across age, sex, smoking, and BMI strata, with no statistically significant interactions detected. Sensitivity analyses also supported the robustness of the primary findings: exclusion of participants with baseline diabetes and complete-case analyses yielded similar overall patterns, and the three-class trajectory solution supported the overall adverse-versus-favorable trajectory pattern. However, because the declining and recovering patterns were not separated in the three-class solution, that analysis could not assess whether recovery after decline carried a distinct risk profile.

This study has several strengths. The use of two large, internationally harmonized prospective cohorts enhances the robustness and external relevance of the findings. A key strength is the direct comparison of cumulative exposure and trajectory pattern within the same analytic framework, which allowed assessment of whether these longitudinal summaries convey overlapping or distinct prognostic information. In addition, prespecified LCMM model selection criteria improved methodological transparency, multiple imputation reduced bias from complete-case restriction, and exclusion of eGDR component variables from the adjustment set helped avoid over-adjustment.

Several limitations should be noted. First, CVD ascertainment relied on self-reported physician diagnosis, which may introduce misclassification, particularly for subclinical events, although self-reported disease history has generally shown acceptable reliability in cohort studies.(55–59) Second, trajectory modeling required repeated eGDR measurements across three waves and therefore excluded participants who died or dropped out before trajectory classification, potentially introducing healthy survivor bias. Third, residual confounding from unmeasured factors, such as diet and genetic susceptibility, cannot be excluded. Fourth, changes in antihypertensive or antidiabetic treatment during follow-up may have influenced eGDR values independently of underlying insulin sensitivity. Fifth, the study populations consisted predominantly of White, middle-aged and older adults from Western countries, which may limit generalizability to younger individuals, other ethnic groups, and populations with different metabolic risk profiles. Sixth, the relatively small size of the declining-then- rising class limited precision, especially in ELSA. Finally, although the joint models suggest that trajectory class may capture prognostic information beyond cumulative average eGDR, these observational analyses cannot establish the underlying biological mechanisms with certainty.

## Conclusion

In conclusion, longitudinal eGDR trajectories provide independent and incremental prognostic information for incident CVD beyond traditional risk factors and time-averaged insulin sensitivity. Persistently low or declining trajectories are associated with substantially higher CVD risk, while trajectory recovery, even among individuals with similar cumulative eGDR exposure, is associated with relative cardiovascular protection. These findings support the value of serial eGDR monitoring as a practical, low-burden tool for dynamic cardiovascular risk stratification in middle-aged and older adults. Future research should examine whether interventions that shift individuals toward more favorable eGDR trajectories reduce CVD incidence, and validate these trajectory-based findings in more diverse populations.

## List of abbreviations

aBIC: Sample-size adjusted Bayesian Information Criterion
AIC: Akaike Information Criterion
AUC: Area under the receiver operating characteristic curve
BIC: Bayesian Information Criterion
BMI: Body mass index
CI: Confidence interval
CRP: C-reactive protein
CV: Coefficient of variation
CVD-: Cardiovascular disease
DCA-: Decision curve analysis
eGDR-: Estimated glucose disposal rate
ELSA-: English Longitudinal Study of Ageing
HbA1c-: Glycated haemoglobin
HDL-C-: High-density lipoprotein cholesterol
HOMA-IR-: Homeostatic model assessment of insulin resistance
HR-: Hazard ratio
HRS-: Health and Retirement Study
HT-: Hypertension status
IFCC-: International Federation of Clinical Chemistry
IQR-: Interquartile range
IR-: Insulin resistance
LCMM-: Latent class mixed model
LRT-: Likelihood ratio test
MICE-: Multiple imputation by chained equations
NGSP-: National Glycohemoglobin Standardization Program
Q1–Q4-: Quartiles 1–4
SD-: Standard deviation
TC-: Total cholesterol
TyG-: Triglyceride-glucose index
VIM-: Variability independent of the mean
WC-: Waist circumference

## Data Availability

The data that support the findings of this study are available from Health and Retirement Study (HRS) and English Longitudinal Study of Ageing (ELSA). Restrictions apply to the availability of these data, which were used under license for this study. Data are available at (https://hrs.isr.umich.edu) and (https://www.elsa-project.ac.uk) with the permission of the respective data providers.

https://hrs.isr.umich.edu

## Acknowledgements

The HRS (Health and Retirement Study) is sponsored by the National Institute on Aging (grant number NIA U01AG009740) and is conducted by the University of Michigan. The English Longitudinal Study of Ageing (ELSA) was developed by a team of researchers based at University College London, the Institute for Fiscal Studies, and the National Centre for Social Research. The data were collected by the National Centre for Social Research. The funding is provided by the National Institute on Aging (grant number NIA R01AG017644) and a consortium of UK government departments coordinated by the Economic and Social Research Council (ESRC). The ELSA data are deposited with the UK Data Service. The authors also thank all participants of both cohorts who contributed their data.

## Declarations

The authors declare no competing interests.

## Ethics approval and consent to participate

The Health and Retirement Study was approved by the University of Michigan Health Sciences and Behavioral Sciences Institutional Review Board (Protocol No. HUM00061128). The English Longitudinal Study of Ageing received ethical approval from the relevant UK Research Ethics Committees across waves (Wave 2: MREC/04/2/006; Wave 4: 07/H0716/48; Wave 6: 11/SC/0374). All participants provided written informed consent prior to participation. The present study involved secondary analysis of de-identified publicly available data and required no additional institutional ethical review. All procedures performed in studies involving human participants were in accordance with the ethical standards of the institutional and national research committees and with the 1964 Helsinki Declaration and its later amendments or comparable ethical standards.

## Author contributions

All the authors contributed substantially to the completion of this study. RP and ML contributed to the conception and design of the study. XL, JZ and CZ contributed to data acquisition and cleaning. RP, XCL and ZW contributed to data analysis and drafted the manuscript. WH, LM and ML critically revised the manuscript. All authors read and approved the final manuscript and agree to be accountable for all aspects of the work.

## Contributor Information

Mingyan Li

## Supplemental material

Figure S1-S14 Table S1-S14

